# Genetic predisposition to high BMI and changes in BMI during adolescence modulate associations between adult BMI and plasma molecules involved in glucose metabolism

**DOI:** 10.1101/2025.09.23.25336458

**Authors:** Alvaro Obeso Fernandez, Gabin Drouard, Aline Jelenkovic, Sari Aaltonen, Teemu Palviainen, Katja M. Kanninen, Jessica E. Salvatore, Danielle M. Dick, Karri Silventoinen, Jaakko Kaprio

## Abstract

**Introduction:** Glucose metabolism and related blood molecules are strongly associated with adult body mass index (BMI). However, it is unclear whether the magnitude of these associations differs in people whose BMI increased rapidly during adolescence or who have a genetic predisposition to high BMI.

**Methods:** The analyses included 373 Finnish participants (39% males) with BMI data at approximate ages 11.5, 14, 17.5, 22, and 37 years. BMI trajectories (defined as BMI changes (i.e. slope) and BMI baseline (i.e. intercept)) were calculated during adolescence (from 11.5 to 17.5 years of age) using linear mixed-effects (LME) models and adulthood (from ∼22 to 37 years of age). We employed LME models to quantify the associations between BMI trajectories during adulthood and nine plasma molecules related to glucose metabolism measured at age ∼22 years. For significant associations, we tested whether the levels of the molecules interacted with either changes in BMI during adolescence or a Polygenic Risk Score (PRS) of BMI (PRS_BMI_). Stratified analyses were performed for men and women.

**Results:** During adulthood, BMI changes per year were 0.16 kg/m^2^ in men and 0.17 kg/m^2^ in women. The analysis revealed seven statistically significant associations between plasma molecules with adult BMI, but no associations with weight change during adulthood. Four statistically significant interactions with plasma molecules were identified, all of which were found in women. In predicting BMI at ∼22 years old, BMI changes in adolescence interacted positively with glucose (p=2.0e-04) and negatively with citrate levels (p= 5.6e-04), and the PRS_BMI_ interacted positively with glucose (p= 3.4e-04) and negatively with citrate (p= 8.3e-03).

**Conclusion:** The current study provides evidence that PRS_BMI_ and BMI changes during adolescence modulate the associations between BMI during adulthood with glucose and citrate blood levels in women.

## Introduction

According to the World Health Organization (WHO), the prevalence of obesity has nearly tripled in the last 40 years (1). This equates to around 3 billion people, including 300 million children and adolescents, who are affected by obesity or are overweight (40% of the global population) (2). Therefore, in order to prevent the increasing prevalence of obesity worldwide, it is crucial to understand the mechanisms that cause or are strongly associated with obesity. A substantial body of research has demonstrated a robust correlation between glucose metabolism and obesity, as well as body mass index (BMI) (3,4). For instance, the relationship between obesity, insulin resistance, imbalance in glucose metabolism and increased fat storage in different tissue types (the main cause of obesity, the excess accumulation of fat in different tissues) has been previously reported by numerous studies (5, 6, 7,8).

The associations between obesity in childhood, adolescence, and adulthood have been intensively studied. Previous studies have indicated that there is an increased risk of developing obesity in adulthood related to weight and obesity status in childhood and adolescence (9, 10). Furthermore, previous studies have demonstrated that there is strong continuity of BMI over childhood (11), indicating that having a high BMI during childhood is an important risk factor for the development of adult obesity (12). Other studies have demonstrated a strong correlation between the plasma proteome and metabolome and changes in BMI (13, 14, 15, 16). Therefore, these studies indicate that alterations in BMI have an impact on an individual’s blood plasma profile. Consequently, it may be hypothesised that the associations between BMI and plasma glucose metabolism molecules could be influenced by past changes in BMI.

Genetic factors are a significant contributor to inter-individual variability in BMI and the associations between glucose metabolism and obesity (17). Previous twin studies have demonstrated that genetic factors account for approximately 75% of the variation in BMI (18). Genome-wide association (GWA) studies have identified numerous genetic variants that contribute to inter-individual differences in BMI during childhood (19) and adulthood (20). More recently, observational studies examining mid- or long-term changes in BMI have shown strong associations between weight trajectories and polygenic risk scores for obesity (21) and blood plasma omics (22).

Glucose metabolism is a central biochemical pathway providing energy and carbon building blocks to growing cells. Different type of molecules take part in this processes such as metabolites, enzymes, coenzymes, nucleosides among others. All this molecules can be found in different steps of the pathway. The molecules used in the current study can be divided in two different groups: enzymes and metabolites. In the first group, we included: Fructose Biphosphate aldolase A and B (FBPALD.A and FBPALD.B), Lactate dehydrogenase A and B (L-LDH.A and L-LDH.B) and Glyceraldehyde 3-phosphate dehydrogenase (GAPDH). In the second group, the following molecules were included: Glucose, lactate, pyruvate and citrate. FBPALD.A and FBPALD.B are a glycolytic enzymes that catalyse the conversion of fructose 1-6-diphosphate to glyceraldehyde 3-phosphate and dihydroxy-acetone phosphate via the glycolysis metabolic pathway (23). Secondly, L-LDH.A and L-LDH.B catalyzes the conversion of pyruvate to lactate and back and at the same time converts NAD+ to NADH and back (24). Lastly, the GAPDH catalyzes the conversion of glyceraldehyde-3-phosphate to 1,3-bisphosphoglycerate as part of glycolysis (25). From the molecules part we have glucose, considered important for being the primary source of energy. Lactate, whose importance lies in providing energy for oxidative metabolism in many tissues and helping maintain redox homeostasis and tissue and whole-organism integrity (26). Pyruvate which is considered an essential molecule being a key molecule in energy production and antioxidant processes apart from being principal connector molecule between processes like glycolysis, gluconeogenesis, fatty acid metabolism, and amino acid metabolism, among others (27). The last metabolite is citrate. This molecule is found in a critical step at a crossroad in intermediary metabolism in humans cells. It acts as a key regulator of energy production inhibiting and inducing strategic enzymes found at the entrance and/or at the exit of different processes such as glycolysis, tricarboxylic acid (TCA) cycle, gluconeogenesis, and fatty acids synthesis (28).

Consequently, it could be hypothesised that genetic factors are also key in explaining how obesity or changes in BMI during adolescence modulate the associations between glycolysis pathway molecule levels. However, to the best of our knowledge, there are no previous studies that have analysed this. Therefore, the aim of this study was to investigate (1) whether BMI or changes in BMI during adulthood are associated with the levels of plasma molecules related to glucose metabolism measured in early adulthood (at ∼22 years of age), and (2) whether genetic predisposition to high BMI and/or changes in BMI during adolescence modulates the identified associations between plasma molecules and BMI or changes in BMI during adulthood.

## Material and methods

### Cohort

The data used in this study was derived from the FinnTwin12 study (FT12). The study includes twins born between 1983 and 1987 with a baseline assessment at age 11.5 and who have been followed up at 14, 17.5, ∼22 and 37 years of age (24, 29, 30). For each of the five surveys, twins completed questionnaires and reported their weight (in kilograms) and height (in centimetres). These data were used to calculate BMI (kg/m^2^). Fasting venous blood samples for a subset of these twins were collected at a mean age of 22 (range 21.0-25.0). In the current analysis, we selected twins who had all five measures of their self-reported BMI as well as blood samples available. Following the exclusion of pregnant women, the final sample comprised 373 twins.). The ethics committee of the Helsinki University Central Hospital District (HUS) approved the most recent data collection (wave 5) (HUS/2226/2021, dated September 22, 2021), and the use of prior collected data. The participants give their informed consent in each wave.

### Quantification of plasma molecules

The plasma molecules analyzed in this study were proteins FBPALD. A & FBPALD. B, L-LDH.A & L-LDH.B and GAPDH), and metabolites (glucose, pyruvate, citrate and lactate). The aforementioned molecules were quantified independently from the same plasma samples of FinnTwin12 participants. This selection was based on our interest in molecules involved in glucose metabolism, with the exclusion of other measured molecules from the analyses, the descriptions of which are available elsewhere (24).

A total of 786 FinnTwin12 participants underwent an in-person clinical visit at approximately 22 years of age, during which their plasma samples were initially quantified for hundreds of proteins. The proteins were processed according to the standard protocol of the Turku Proteomics Facility (Turku, Finland), which involved precipitation and in-solution digestion. This procedure was conducted in accordance with previously published methods (31). The samples were initially analyzed by independent data acquisition LC-MS/MS using a Q Exactive HF mass spectrometer, and subsequently analyzed using Spectronaut software.The raw proteomic data was subjected to local normalization (32), processing, and quality control in accordance with the methodology described elsewhere (33). In brief, protein levels were transformed using the log_2_ function and proteins with >10% missing values were excluded. Missing values were imputed using the lowest observed value for each protein. Correction for batch effects was performed using Combat (33). From this dataset, the proteins FBPALD.B, FBALD.A, L-LDH.A, L-LDH.B and GAPDH were selected, and their levels were scaled to have a mean of zero and a variance of one.

Metabolomic data was quantified from plasma samples of FinnTwin12 participants using high-throughput proton nuclear magnetic resonance spectroscopy (1H-NMR) (Nightingale Health Ltd, Helsinki, Finland) (29, 34, 35), as previously described in detail elsewhere (36). The four metabolites of interest (glucose, lactate, pyruvate and citrate) were derived from a dataset comprising 140 variables (37). The data were processed in accordance with the following steps: exclusion of pregnant women and individuals on cholesterol medication, imputation of missing values using the observed minimum sample value, and outlier checking. Metabolite values were subsequently normalized using inverse normal rank transformation.

### Polygenic Risk Score of Obesity

A polygenic risk score (PRS) of BMI (PRS_BMI_) was calculated to test for interactions with plasma molecules in predicting BMI and/or changes in BMI during adulthood Genotyping was performed using Illumina Human610-Quad v1.0 B, Human670-QuadCustom v1.0 A, Illumina HumanCoreExome (12 v1.0 A, 12 v1.1 A, 24 v1.0 A, 24 v1.1 A, 24 v1.2 A) and Affymetrix FinnGen Axiom arrays. The technical details of genotyping and imputation have been previously described elsewhere (38). Polygenic risk score for BMI were calculated using summary statistics from Yengo et al. (2018) having weights available for total of 692,578 individuals and implementing a Bayesian approach taking account the linkage disequilibrium between each SNP by re-weighting the GWAS summary statistics using an external LD reference panel. The LD reference panel were consisting of 27284 unrelated Finnish samples from the national FINRISK study (39). The infinitesimal model with default parameters were applied using LDpred for re-weighting and polygenic score calculation (40). The SNPs for polygenic scoring were selected by following the guidance to choose a set of SNPs representing the whole genome which has been pruned to an amount that will keep the computational burden reasonable. Therefore, we choose to use ∼1 million HapMap3 SNPs (The actual SNP list can be downloaded from https://csg.sph.umich.edu/abecasis/mach/download/HapMap3.r2.b36.html) with minor allele frequency >5% in European individuals with major histocompatibility complex region from chromosome 6 (GRCh37: 6p22.1-21.3) excluded (41). After these steps,a total of 996,919 common single nucleotide polymorphisms (SNPs) were used for final polygenic scoring. A regression analysis was conducted to correct for population stratification, with the PRS regressed on the top ten genetic principal components (42). The residuals were then scaled to a mean of zero and unit variance, and the PRS was used unmodified in relevant analyses.

In the present study, the PRS values were classified into three distinct groups based on their normal distribution. The first group included individuals with PRS values less than -1, who were considered to have a low genetic predisposition to a high BMI. The second group consisted of individuals with PRS values between -1 and 1, who were considered to have a normal genetic predisposition to a high BMI. The third and final group included individuals with PRS values greater than 1, who were considered to have a high genetic predisposition to a high BMI.

### Calculation of changes in BMI during adolescence and adulthood

The BMI measurements were divided into two groups. The first group included BMI measures corresponding to adolescence (11.5, 14 and 17.5 years), while the second group included measurements during adulthood (∼22 and 37 years). To calculate the changes in BMI during adolescence, we used linear mixed-effects (LME) modelling, which incorporates both fixed and random effects (40). These models have been used extensively in the literature to model BMI trajectories (41, 42) as they allow researchers to extract intercept and slope values for each individual, representing baseline BMI and rates of change in BMI. We therefore fitted LME models to derive intercept and slope values for each individual, using a random effect on individual identifiers. For the change in BMI during adulthood, we used the delta method, which subtracts BMI values at average ages 22 and 37 and then divides by the time elapsed between the two measurements. In both cases, the changes in BMI during adolescence and adulthood were expressed in kg/m^2^ per year.

### Association and interaction analyses

The present study employed LME models to investigate the associations between body mass index (BMI) and changes in BMI during adulthood with plasma molecule levels. The use of LME models, as opposed to more classical approaches such as linear regression models, was motivated by the correlation structure of the data, in that family relatedness violates the assumption of independence of observations in classical linear regression models, which we corrected by using LME models with random effects on family identifiers. Furthermore, interactions between plasma molecule levels with PRSBMI and changes in BMI during adolescence were tested. First, to asses if the sex differentiations in BMI levels reported in the descriptive information displayed in Table 1 were statisticaly significant we fitted Model 1.

**Table 1.**
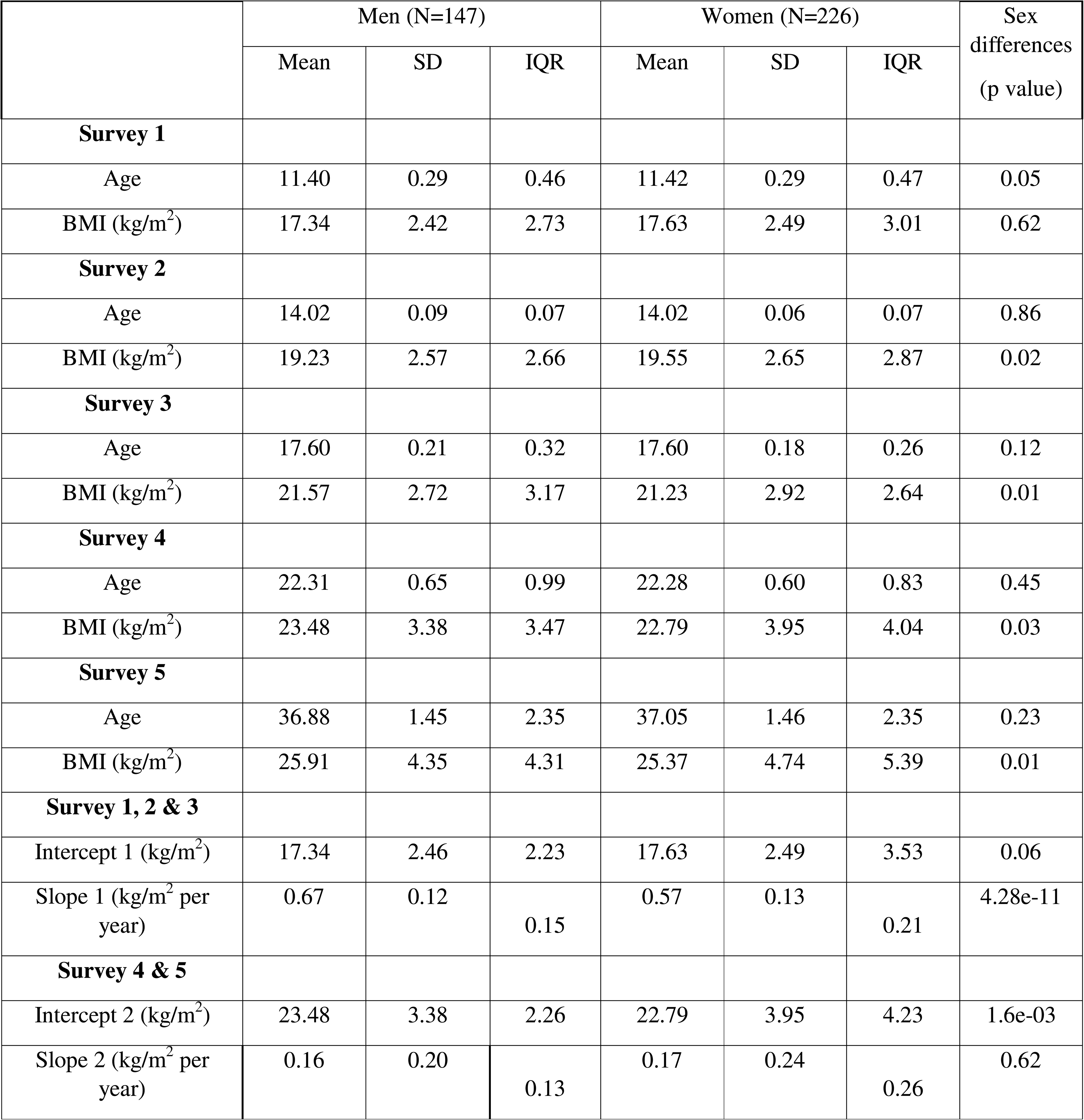
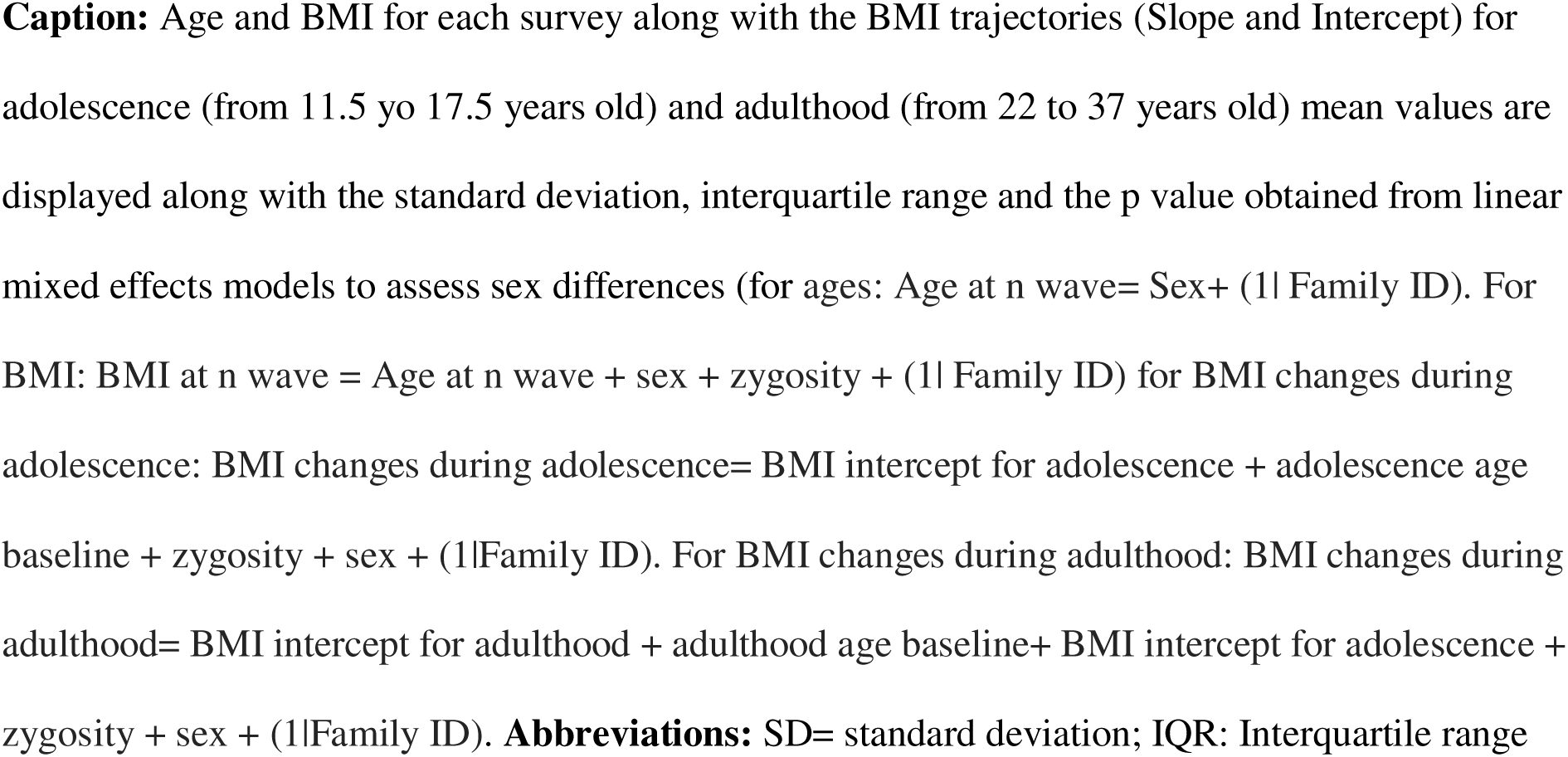
Descriptive information of FinnTwin12 participants by sex.

Model 1: Change_BMIadolescence OR BMI22 ∼ age at blood sample + BMI_baseline + sex+ (1|Family ID)

Once the differences were demonstrated to be statistically significant, appart from the well-known differences in glucose metabolism between the sexes (43, 44), separate LME models were fitted for males and females,proceding to run Model 2. This was fitted to quantify associations between changes in BMI during adulthood or BMI levels at ∼22 and 37 years of age with plasma molecule levels.

**Model 2:** Change_BMI_adulthood_ *OR* BMI_22_ *OR* BMI_37_ ∼ age at blood sample + molecule + (1|Family ID)

Secondly, significant associations were subjected to interaction analyses. The aim of this study was to assess whether changes in BMI during adolescence or PRS_BMI_ modulate associations between BMI trajectories during adulthood and plasma molecules. To this end, Model 3 and Model 4 were fitted to the data.

Model 3: Change_BMIadulthood OR BMI∼22 OR BMI37 ∼ age at blood sample + molecule + molecule:Change_BMIadolescence + Change_BMIadolescence + (1|Family ID)

Model 4: Change_BMIadulthood OR BMI∼22 OR BMI37 ∼ age at blood sample + molecule + molecule: PRSBMI + PRSBMI + (1|Family ID)

Finally, two sensitivity analyses were conducted. The first aimed to assess whether significant interactions between changes in BMI during adolescence and plasma molecules decreased when BMI at baseline age of adolescence (i.e., BMI at 11.5 years of age) was added. The second sought to determine whether the associations found at all levels remained significant after individuals with BMI > 30 kg/m2 at ∼22 and 37 years old (15 samples) were removed. In the affirmative case, we sought to quantify the strength of the association. All analyses were performed using the R software (version 4.2.3) and the R package lme4 (version 1.1-34).

## Results

### Trajectories of BMI during adolescence and adulthood

Both men and women showed an increase in BMI from early adolescence to adulthood (from 11.5 to 37 years of age) with a mean BMI of 8.57 kg/m² in men and 7.74 kg/m² in women. The changes in BMI per year were found to be greater during adolescence compared to adulthood. In boys, the change in BMI per year was 0.67 kg/m2, while it was 0.57 kg/ m2 in girls. In contrast, in men, the change in BMI per year was found to be only 0.16 kg/m2, while in women, the change was 0.17 kg/m2.

Males had greater BMI at blood sampling (p=6.6e-9) and greater change in BMI (p=1.3e-15) than females, as assessed with linear mixed-effects models adjusting for age and baseline adolescent BMI. During adolescence, changes in BMI and the BMI at baseline age (i.e., BMI at 11.5 years of age) were weakly positively associated in men and moderately positively associated in women. However, changes in BMI during adulthood and baseline BMI (i.e., BMI at ∼22 years of age) were negatively correlated in both sexes. However, only in women was this correlation significant (Table 2). This suggests that the higher the BMI at ∼ 22 years, the lower the increase in BMI from ∼22 to 37 years in women. Moreover, changes in BMI during adolescence were weakly negatively correlated with changes in BMI during adulthood in men and women. The results suggests that there are differences in the rates of weight gaining or shifting between these two periods of life.

**Table 2:** Pairwise Pearson correlation between BMI measurements and changes in BMI during adolescence or adulthood.

|  | Men |  |  |  | Women |  |  |  |
| --- | --- | --- | --- | --- | --- | --- | --- | --- |
|  | Coefficient | 95% CI |  | p value | Coefficient | 95% CI |  | p value |
|  |  | LB | UB |  |  | LB | UB |  |
| **Adolescence (ages 11.5 to 17.5)** |  |  |  |  |  |  |  |  |
| Slope 1 vs Intercept 1 | 0.22 | 0.06 | 0.36 | 7.05e-03 | 0.38 | 0.26 | 0.48 | 3.85e-09 |
| **Adulthood (ages ~22 to 37)** |  |  |  |  |  |  |  |  |
| Slope 2 vs Intercept 2 | -0.07 | -0.22 | 0.09 | 0.39 | -0.20 | -0.32 | -0.07 | 2.25e-03 |
| **Adolescence and adulthood** |  |  |  |  |  |  |  |  |
| Slope 1 vs Slope 2 | -0.16 | -0.31 | -0.01 | 0.05 | -0.12 | -0.25 | -0.01 | 0.05 |

**Caption:** Pairwise phenotypic correlations between the changes in BMI during adolescence (i.e. slope 1) (from 11.5 to 17.5 years old) and adulthood (i.e. slope 2) (from 22 to 37 years old) along with the BMI baseline values at adolescence (i.e. Intercept 1) and adulthood (i.e. Intercept 2).
**Abbreviations:** LB: Lower bound; UB: Upper bound; CI: Confidence interval.

Associations between BMI trajectories and plasma molecules without interactions In men, we observed an association between BMI in adulthood and two specific molecules. Firstly, FBPALD.B was found to be positively associated with BMI at ∼22 years of age and at 37 years of age. Secondly, glucose was positively associated with BMI at 37 years of age (Table 3).

**Table 3:** Linear mixed-effects models quantifying associations between plasma molecules (measured in standard deviation units) of interest with BMI and changes in BMI during adulthood (i.e., slope 2) (from 22 to 37 years old) by sex.

| Metabolite<br>(SD) | Slope 2 (kg/m2 per year) |  |  |  | BMI at ~22 (kg/m2) |  |  |  | BMI at 37 (kg/m2) |  |  |  |
| --- | --- | --- | --- | --- | --- | --- | --- | --- | --- | --- | --- | --- |
|  | Value | 95% CI |  | p value | Value | 95% CI |  | p value | Value | 95% CI |  | p value |
|  |  | LB | UB |  |  | LB | UB |  |  | LB | UB |  |
| **Men** |  |  |  |  |  |  |  |  |  |  |  |  |
| FBPALD.A | -0.01 | -0.04 | 0.02 | 0.57 | -0.07 | -0.65 | 0.49 | 0.79 | -0.23 | -0.95 | 0.47 | 0.52 |
| FBPALD.B | 0.01 | -0.03 | 0.04 | 0.77 | 0.70 | 0.17 | 1.25 | 7.15e-03 | 0.73 | 0.08 | 1.39 | 0.02 |
| GAPDH | -0.02 | -0.05 | 0.01 | 0.22 | 0.01 | -0.45 | 0.45 | 0.97 | -0.28 | -0.83 | 0.25 | 0.30 |
| L-LDH.A | 0.01 | -0.03 | 0.04 | 0.73 | 0.40 | -0.12 | 0.93 | 0.13 | 0.37 | -0.28 | 1.06 | 0.26 |
| L-LDH.B | -0.02 | -0.05 | 0.01 | 0.28 | 0.07 | -0.44 | 0.59 | 0.77 | -0.20 | -0.84 | 0.44 | 0.54 |
| Glucose | 0.02 | -0.01 | 0.05 | 0.33 | 0.26 | -0.26 | 0.78 | 0.33 | 0.65 | 1.24e-03 | 1.30 | 0.05 |
| Lactate | -0.02 | -0.05 | 0.01 | 0.34 | -0.17 | -0.73 | 0.36 | 0.51 | -0.43 | -1.10 | 0.23 | 0.20 |
| Citrate | 0.01 | -0.02 | 0.04 | 0.47 | 0.10 | -0.40 | 0.60 | 0.69 | 0.28 | -0.36 | 0.93 | 0.38 |
| Pyruvate | 0.01 | -0.02 | 0.04 | 0.55 | -0.22 | -0.72 | 0.27 | 0.38 | -1.20e-04 | -0.63 | 0.63 | 1 |
| **Women** |  |  |  |  |  |  |  |  |  |  |  |  |
| FBPALD.A | -0.01 | -0.04 | 0.01 | 0.31 | 0.19 | -0.26 | 0.64 | 0.41 | -0.03 | -0.61 | 0.54 | 0.86 |
| FBPALD.B | -0.01 | -0.04 | 0.01 | 0.33 | 0.20 | -0.21 | 0.62 | 0.34 | 0.07 | -0.46 | 0.60 | 0.79 |
| GAPDH | 0.01 | -0.02 | 0.04 | 0.57 | -0.38 | -0.82 | 0.06 | 0.09 | -0.26 | -0.80 | 0.30 | 0.36 |
| L-LDH.A | -0.01 | -0.04 | 0.02 | 0.55 | 0.32 | -0.11 | 0.76 | 0.14 | 0.24 | -0.30 | 0.80 | 0.39 |
| L-LDH.B | 3.15e-03 | -0.03 | 0.02 | 0.83 | 0.02 | -0.44 | 0.48 | 0.93 | -0.07 | -0.66 | 0.52 | 0.81 |
| Glucose | -0.02 | -0.05 | 0.01 | 0.19 | 0.73 | 0.17 | 1.28 | 0.01 | 0.50 | -0.20 | 1.20 | 0.16 |
| Lactate | 0.01 | -0.02 | 0.04 | 0.54 | 0.13 | -0.43 | 0.68 | 0.64 | 0.47 | -0.25 | 1.17 | 0.17 |
| Citrate | 7.12e-03 | -0.04 | 0.02 | 0.66 | -1.02 | -1.55 | -0.49 | 1.80e-04 | -1.15 | -1.82 | -0.49 | 7.07e-04 |
| Pyruvate | -0.01 | -0.04 | 0.02 | 0.51 | 0.57 | 0.06 | 1.08 | 0.02 | 0.41 | -0.22 | 1.05 | 0.20 |

**Caption:** Results are displayed in males and females separately. The model used to quantify associations was model 1 (see Methods), with the changes in BMI or BMI during adulthood (for changes in BMI the age range is 22 to 37 years) as an outcome, and the molecule and the age at blood sampling as fixed effects. **Abbreviations:** LB: Lower bound; UB: Upper bound; SD: standard deviation; FBPALD.A: Fructose bisphosphate aldolase subunit A; FBPALD.B: Fructose bisphosphate aldolase subunit B; GAPDH: Glyceraldehyde 3-phosphate dehydrogenase; L.LACTDH.A: L- Lactate dehydrogenase subunit A; L.LACTDH.B: L- Lactate dehydrogenase subunit ; BMI: Body mass index

In women, blood glucose, citrate, and pyruvate concentrations were significantly associated with BMI measurements. First, glucose was positively associated with BMI at ∼22 years of age. Moreover, citrate was found to be negatively associated with both BMI at ∼22 and 37 years of age. Finally, pyruvate showed positive association with BMI at ∼22 years of age (Table 3).

Sensitivity analyses, conducted by removing individuals with BMI>30 kg/m^2^, resulted in the maintenance of the previously significant associations, although the value of the estimate decreased by 40-50%.

### Associations between BMI trajectories and plasma molecules with interactions

Significant results were obtained in four situations, all of which involved women, after conducting models with BMI at ∼22 and 37 years as an outcome and with the plasma molecules that showed a significant association in the previous section. The results of these four interactions are displayed in Figure 1 and Table 4.

**Figure 1:**
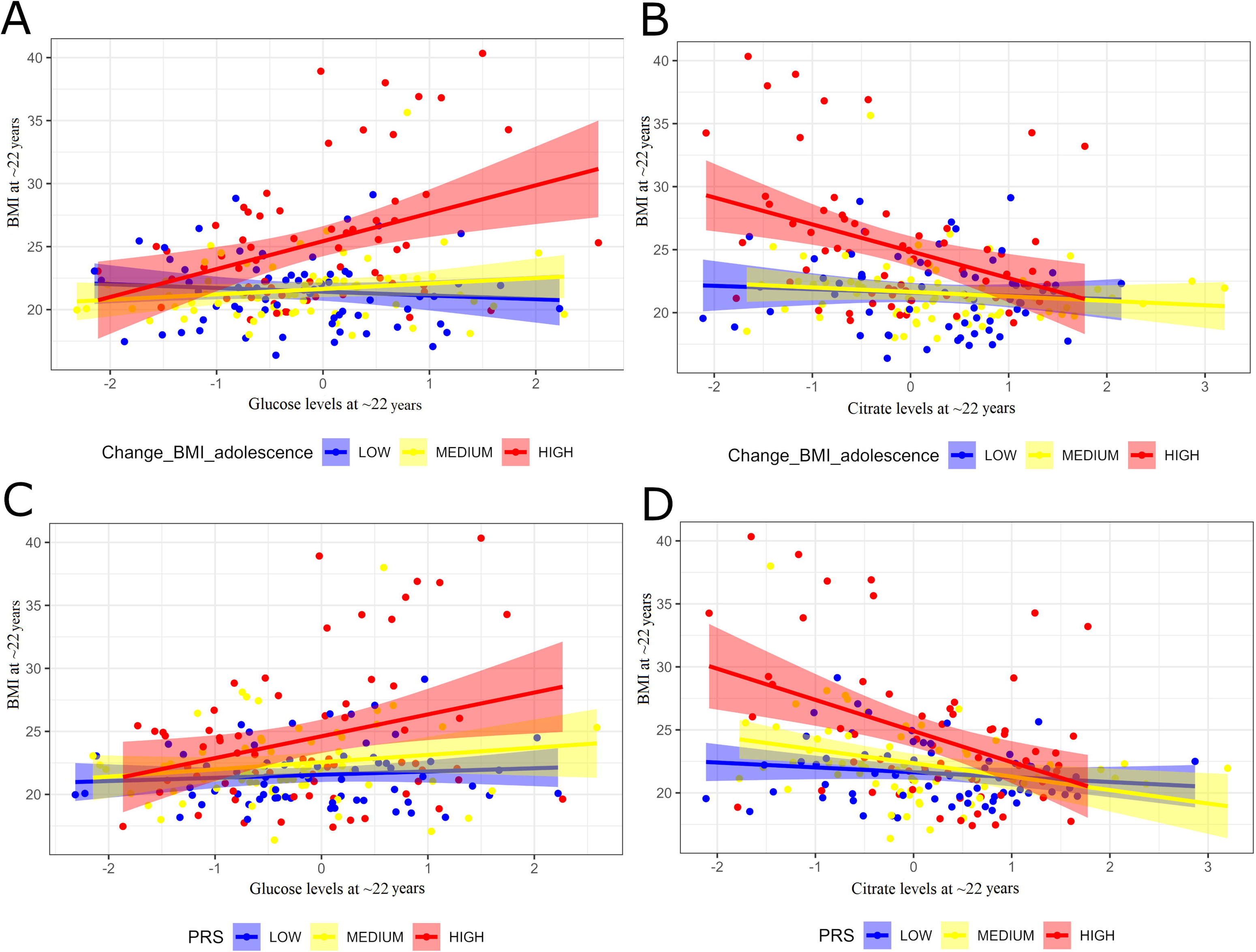
Graphical illustration of interactions between plasma molecule levels with changes in BMI during adolescence or the PRS of BMI in predicting BMI levels during adulthood. **Caption: (A)** Graphical illustration of the association between BMI levels at ∼22 years old and the interaction between glucose and BMI changes in adolescence in women (interaction coefficient: 6.08, p=2.01e-04). The changes in BMI were divided into three categories after scaling the values to mean zero and variance one: low (below 1 SD or 3,95 kg/m^2^ from the mean), high (greater than 1 SD or 3,95 kg/m^2^ from the mean), and medium (within 1 SD or 3,95 kg/m^2^ from the mean). **(B)** Graphical illustration of the association between BMI levels at ∼22 years old and the interaction between citrate and BMI changes in adolescence in women (interaction coefficient: -5.46, p=5.59e-04). The changes in BMI were divided into three categories after scaling the values to mean zero and variance one: low (below 1 SD or 3,95 kg/m^2^ from the mean), high (greater than 1 SD or 3,95 kg/m^2^ from the mean), and medium (within 1 SD or 3,95 kg/m^2^ from the mean). **(C)** Graphical illustration of the association between BMI levels at ∼22 years old and the interaction between glucose and the polygenic risk score of BMI (PRS_BMI_) in women. The PRS of BMI is divided into three categories after scaling the values to mean zero and variance one: low (below 1 SD from the mean), high (greater than 1 SD from the mean), and medium (within 1 SD from the mean). **(D)** Graphical illustration of the association between BMI levels at ∼22 years old and the interaction between citrate and the polygenic risk score of BMI in women. The PRS_BMI_ is divided into three categories after scaling the values to mean zero and variance one: low (below 1 SD from the mean), high (greater than 1 SD from the mean), and medium (within 1 SD from the mean). Abbreviations: BMI: Body mass index. SD: standard deviation.

**Table 4:** Linear mixed-effects models to assess interactions between plasma molecules (measured in standard deviation units) with BMI changes in adolescence (i.e. Slope 1) or the genetic predisposition to BMI (PRS_BMI_).

| Outcome | Metabolite<br>(SD) | Slope 1 (kg/m2 per year) |  |  |  | Metabolite (SD) |  |  |  | Metabolite:Slope 1 |  |  |  |
| --- | --- | --- | --- | --- | --- | --- | --- | --- | --- | --- | --- | --- | --- |
|  |  | Value | 95% CI |  | p value | Value | 95% CI |  | p value | Value | 95% CI |  | p value |
|  |  |  | LB | UB |  |  | LB | UB |  |  | LB | UB |  |
| **Men** |  |  |  |  |  |  |  |  |  |  |  |  |  |
| BMI at ~22 (kg/m2) | FBPALD.B | 11.19 | 7.23 | 15.26 | 2.84e-07 | -0.18 | -2.68 | 2.33 | 0.88 | 0.77 | -2.67 | 4.22 | 0.66 |
| BMI at 37 (kg/m2) | FBPALD.B | 6.28 | 1.01 | 11.7 | 0.02 | 0.47 | -2.93 | 3.93 | 0.78 | 0.08 | -4.63 | 4.73 | 0.97 |
| BMI at 37 (kg/m2) | Glucose | 8.68 | 3.74 | 13.77 | 7.04e-04 | 3.82 | 0.57 | 7.14 | 0.02 | -4.28 | -9.05 | 0.37 | 0.07 |
| **Women** |  |  |  |  |  |  |  |  |  |  |  |  |  |
| BMI at ~22 (kg/m2) | Glucose | 15.05 | 11.50 | 18.58 | 2.16e-15 | -3.00 | -4.82 | -1.17 | 1.62e-03 | 6.08 | 2.96 | 9.19 | 2.01e-04 |
| BMI at ~22 (kg/m2) | Citrate | 15.53 | 12.10 | 18.97 | <2.20e-16 | 2.54 | 0.68 | 4.38 | 8.04e-03 | -5.46 | -8.49 | -2.42 | 5.59e-04 |
| BMI at ~22 (kg/m2) | Pyruvate | 16.47 | 12.99 | 19.97 | <2.20e-16 | 0.15 | -1.79 | 2.10 | 0.87 | 0.52 | -2.67 | 3.73 | 0.74 |
| BMI at 37 (kg/m2) | Citrate | 11.81 | 6.99 | 16.64 | 1.67e-06 | 1.24 | -1.36 | 3.88 | 0.35 | -3.65 | -8.01 | 0.63 | 0.09 |
|  |  | PRS<sub>BMI</sub> |  |  |  | Metabolite (SD) |  |  |  | Metabolite:PRS<sub>BMI</sub> |  |  |  |
| **Men** |  |  |  |  |  |  |  |  |  |  |  |  |  |
| BMI at ~22 (kg/m2) | FBPALD.B | 0.69 | 0.12 | 1.26 | 0.01 | 0.73 | 0.20 | 1.28 | 6.90e-03 | 0.09 | -0.46 | 0.65 | 0.73 |
| BMI at 37 (kg/m2) | FBPALD.B | 1.42 | 0.74 | 2.10 | 7.70e-05 | 0.73 | 0.10 | 1.36 | 0.02 | 0.09 | -0.57 | 0.76 | 0.78 |
| BMI at 37 (kg/m2) | Glucose | 1.22 | 0.52 | 1.92 | 9.70e-04 | 0.54 | -0.09 | 1.18 | 0.09 | 0.40 | -0.23 | 1.04 | 0.22 |
| **Women** |  |  |  |  |  |  |  |  |  |  |  |  |  |
| BMI at ~22 (kg/m2) | Glucose | 1.45 | 0.86 | 2.04 | 3.20e-06 | 0.86 | 0.32 | 1.39 | 2.01e-03 | 1.24 | 0.56 | 1.90 | 3.41e-04 |
| BMI at ~22 (kg/m2) | Citrate | 1.34 | 0.75 | 1.93 | 1.20e-05 | -0.97 | -1.5 | -0.45 | 3.16e-04 | -0.74 | -1.30 | -0.19 | 8.34e-03 |
| BMI at ~22 (kg/m2) | Pyruvate | 1.38 | 0.77 | 2.00 | 1.53e-05 | 0.67 | 0.15 | 1.19 | 0.01 | 0.05 | -0.52 | 0.63 | 0.86 |
| BMI at 37 (kg/m2) | Citrate | 1.18 | 0.43 | 1.93 | 1.96e-03 | -1.11 | -1.77 | -0.45 | 1.16e-03 | -0.42 | -1.14 | 0.28 | 0.22 |

**Caption:** Model outputs are displayed in males and females separately. The models fitted were specified as model 2 and model 3 (see Methods), where changes in BMI during adulthood (from 22 to 37 years old) or the BMI at 22 and 37 years old are the outcomes. The molecule level at blood sampling, the age at blood sampling, as well as the changes in BMI during adolescence (model 2) or the PRS of BMI (model 3) were added as fixed effects, in addition to the interaction term between the plasma molecule with either the changes in BMI during adolescence (model 2) or the PRS of BMI (model 3). **Abbreviations:** LB: Lower bound; UB: Upper bound; SD: standard deviation; FBPA.LD.B: Fructose bisphosphate aldolase subunit B; BMI: Body mass index; PRS: Polygenic risk score.

First, we conducted an analysis of the changes in BMI during adolescence and their interaction with the levels of molecules at ∼22 years old. The results indicated that BMI at ∼22 years old was negatively associated with glucose levels (p=2.16e-15) and positively associated with changes in BMI during adolescence (p=2.16e-15), Additionally, the interaction between BMI changes during adolescence and glucose levels was found to be positive (p=2.01e-04) (Table 4; Figure 1a). Furthermore, the BMI at ∼22 years old was found to be positively associated with citrate levels (p=8.04e-03) and changes in BMI during adolescence (p= <2e-16), with citrate levels and changes in BMI during adolescence exhibiting a negative interaction (p= 5.59e-04) (Table 4; Figure 1b).

When conducting a sensitivity analysis by adding the BMI at baseline age from adolescence (i.e., BMI at 11.5 years old) as a covariate in analyses investigating interactions with changes in BMI during adolescence, we found that these interactions did not remain significant (see Supplementary Table 1). This suggests that the interaction between changes in BMI during adolescence with molecules is likely to reflect the effects of adolescence BMI on associations, rather than the change in BMI during adolescence itself. Moreover, after conducting sensitivity analyses following the removal of individuals with BMI>30 kg/m^2^ the previously found significant associations remained, despite the estimated values decreasing by 40 – 50%.

Similarly, we quantified interactions between PRS_BMI_ and molecule levels at ∼22 years of age. The results indicated that there was a positive interaction between glucose and the PRS_BMI_ predicting BMI levels at ∼22 years of age (p= 3.4e-04). This interaction was in addition to the significant individual effects observed (Table 4; Figure 1c). Additionally, BMI at ∼22 years of age was positively associated with the PRS_BMI_ (p=1.2e-05) and negatively associated with citrate levels (p= 3.1e-04), along with a statistically significant negative interaction between citrate and PRS_BMI_ (p= 8.3e-03) (Table 4; Figure 1d).

## Discussion

The present study sought to assess whether changes in BMI during adulthood are associated with 279 the levels of plasma molecules involved in glucose metabolism measured in young adulthood (i.e., at ∼ 22 years of age). Secondly, we sought to investigate whether genetic predisposition to high 281 BMI and/or changes in BMI during adolescence modulate the identified associations between metabolites and changes in BMI during adulthood. We found associations between several glucose metabolism molecules and BMI in adulthood, as well as interactions between molecule levels and either changes in BMI during adolescence or genetic predisposition to high BMI. Interestingly, no associations were observed with changes in BMI during adulthood. In men, a positive association was observed between FBPALD.B and glucose levels and changes in BMI during adulthood. However, no association with interactions between molecule levels and either changes in BMI during adolescence or genetic predisposition to high BMI was found. In women, significant associations were observed between glucose, pyruvate and citrate and BMI in adulthood. The first two associations were positive, while the third was negative. In contrast to the findings in men, women showed 2 significant interactions between molecule levels (glucose and citrate) and changes in BMI during adolescence in explaining variability in BMI in adulthood. Two interactions between plasma molecules levels (glucose and citrate) and the genetic predisposition to high BMI were also identified in women.

Even though it is known in the literature how 1) the glucose metabolism correlates with obesity and BMI, 2) how changes in BMI associates with an individual’s blood plasma profile, and 3) how changes in BMI associate with PRS for obesity and blood plasma omics, there is a lack of knowledge about how associations between BMI and plasma glucose metabolism molecules could be influenced by past changes in BMI and the role played by genetic factor in explaining how obesity or changes in BMI during adolescence modulate the associations between glycolysis pathway molecule levels and BMI. In light of the above considerations, our study provides new results and a deeper understanding of how earlier BMI trajectories in adolescence and genetics modulate associations between glucose metabolism and BMI in adulthood.

We found a positive association of FBPALD.B protein and glucose levels with BMI and its changes in adult men. The higher the levels of each of these molecules, the higher the BMI and its changes in adulthood (∼22 and 37 years of age). In women, we found that glucose and pyruvate exhibited a similar direction of association with BMI in adulthood, whereas citrate exhibited an opposite direction of association. This suggests that elevated levels of citrate were associated with a lower BMI during adulthood. Previous studies have reported an association between glucose and changes in BMI (28, 45), which is particularly relevant given the link between this relationship and the early stages of type 2 diabetes (T2D), where insulin resistance results in increased levels of glucose in the blood (46). With regard to FBPALD.B, it has previously been reported that elevated *ALDOB* gene expression (the gene that encodes the FBPALD.B) in the liver is negatively associated with insulin secretion (47). Consequently, elevated levels of FBPALD.B are associated with reduced insulin secretion, which may exacerbate hyperglycaemia and contribute to the pathogenesis of T2D. Additionally, a candidate gene study by Daimon et al. (2003) found that *ALDOB* (rs506571) is linked to T2D. Regarding citrate molecules, it has been shown that children with obesity have lower levels of citrate than those of a healthy weight (49). Another study showed that in general population, regardless of age, plasma citrate levels are typically elevated in obesity (50). Furthermore, previously conducted research has indicated that there is a strong and significant negative association between the activity of citrate synthase (the enzyme that catalyses the synthesis of citrate) and obesity condition (51). This suggests that as obesity levels increase, the activity of the enzyme and thus the production of citrate is reduced. Finally, with regard to pyruvate, although it has been claimed that the intake of pyruvate significantly enhances weight loss in several intervention studies (52, 53, 54), to the best of our knowledge, we have found no literature referring to any association between this molecule levels in blood and obesity or even gains in BMI.

In the context of interaction analyses, it was found that two molecules (glucose and citrate) interacted with changes in BMI during adolescence to predict BMI in adulthood. For glucose, a significant positive association was observed between BMI at the age of ∼22 years and an interaction with changes in BMI during adolescence. This implies that for two individuals with the same levels of glucose at ∼22 years of age, the one with a higher BMI gain during adolescence had a higher BMI once adult in addition to the individual effect of the change in BMI during adolescence (Table 4; Figure 1a). A previous study proposed that obesity or being overweight during adolescence could result in an impaired glucose profile (higher glucose than normal), and that the level of impairment increased over the years leading to an adulthood obesity (48). Regarding citrate, we found a significant negative association with BMI at ∼22 years old and an interaction with BMI changes during adolescence, meaning that for two individuals with the same BMI gain during adolescence, the one with a greater level of citrate will have a lower BMI during adulthood, in addition to the negative individual effect of citrate alone on BMI (Table 4; Figure 1b). To the best of our knowledge, this result has not been previously reported in the literature.

Finally, in analyses that included interaction terms between the molecules levels and the genetic predisposition to high BMI, we found that glucose and citrate levels interacted with the PRS_BMI_ in predicting BMI at ∼22 years old in women. In the case of glucose, for two individuals with the same PRS_BMI_ value, the individual with a greater level of glucose in plasma will have greater levels of BMI during adulthood in addition to the individual effect of glucose alone (Table 4; Figure 1c). In contrast, with citrate, the opposite occurs: should two individuals have the same PRS_BMI_ value, the one with greater levels of citrate is likely to have lower BMI values during adulthood, in addition to the effect of citrate alone (Table 4; Figure 1d).

The strengths of this study include the use of a cohort of twins with BMI data available from adolescence to mid-adulthood as well as proteomic, metabolomic and PRS data. Nevertheless, it should be noted that the current study is not without limitations. The first limitation is the relatively small sample size. The selection of participants with all BMI measures may have resulted in a reduction of statistical power and increased uncertainty in the estimated outcomes. In addition, relying solely on measures taken at a specific age (in the case of metabolites and proteins) is a limitation as it provides only one isolated data point, rather than multiple measures that would allow for a full longitudinal omic study to be conducted. Moreover, another limitation is the non-inclusion of covariates related to nutrition, such as history of eating/feeding disorders. Finally, although BMI is the most commonly used measurement of obesity, it does not provide information about body composition and thus is less accurate for the measurement of fat mass than if used in conjunction with other measures, such as body fat percentage. However, a change in weight is not solely a change in fat mass.

## Conclusion

Overall, our findings suggest that the associations between BMI or changes in BMI during adulthood and plasma molecules linked to glucose metabolism are subject to modulation in individuals with higher PRS_BMI_ value or greater changes in BMI during adolescence. The obtained results open the door to the development of public health strategies for the prevention of obesity at different periods of life.

## Data Availability Statement

The FT12 data is not publicly available due to the restrictions of informed consent. However, the FT12 data is available through the Institute for Molecular Medicine Finland (FIMM) Data Access Committee (DAC) for authorised researchers who have IRB/ethics approval and an institutionally approved study plan. To ensure the protection of privacy and compliance with national data protection legislation, a data use/transfer agreement is needed, the content and specific clauses of which will depend on the nature of the requested data.

## Human Ethics and Consent to Participate declarations

The Ethics Committee of the Helsinki University Central Hospital District (HUS) approved the most recent data collection (wave 5) (HUS/2226/2021, dated September 22, 2021) as well as the use of previously collected data. The participants or their parents/legal guardians provided informed consent when participating in the surveys.

## Clinical trial number

not applicable.

## Consent to Publish declaration

not applicable.

## Code availability

All R scripts are available from the corresponding authors.

## Acknowledgements

Not applicable.

## Author contributions

The study design was developed and discussed by AO, GD and JK. The statistical analyses were performed by AO. The phenotype data was processed by AO and plasma omic data processing was performed by GD. SA, KMK, JES, DMD, and JK participated in the data collection. Polygenic risk scores were calculated by TP. AO wrote the original manuscript. All authors actively participated in the improvement of the manuscript by critically revising it. All of the authors read and approved the final version of the manuscript.

## Competing interests

The authors declared no potential conflicts of interest with respect to the research, authorship, and/or publication of this article.

## Funding

Phenotype and omics data collection in FinnTwin12 cohort has been supported by the Wellcome Trust Sanger Institute, the Broad Institute, ENGAGE – European Network for Genetic and Genomic Epidemiology, FP7-HEALTH-F4-2007, grant agreement number 201413, National Institute of Alcohol Abuse and Alcoholism (grants AA-12502, AA-00145, and AA-09203 to R J Rose and AA15416 and K02AA018755 to D M Dick, and R01AA015416 to J Salvatore), the Academy of Finland (grants 100499, 205585, 118555, 141054, 264146, 308248 to JK, and grants 328685, 307339, 297908 and 251316 to M Ollikainen, and the Centre of Excellence in Complex Disease Genetics grants 312073, 336823, and 352792 to JK), and Sigrid Juselius Foundation (to MO). This research was partly funded by the European Union’s Horizon 2020 research and innovation program under grant agreement No 874724 (Equal-Life). Equal-Life is part of the European Human Exposome Network. KS and JK have been supported by the European Union’s Horizon Europe Research and Innovation programme under Grant Agreement number 101080117. Views and opinions expressed are however those of the author(s) only and do not necessarily reflect those of the European Union. Neither the European Union nor the granting authority can be held responsible for them.

## Notes

### Competing Interest Statement

The authors have declared no competing interest.

