## Supplementary tables for "Genetic predisposition to high BMI and changes in BMI during adolescence modulate associations between adult BMI and plasma molecules involved in glucose metabolism"

|  |  | Slope 1 (BMI unit/year) | | | | Metabolite (SD) | | | | Metabolite: Slope 1 | | | | Intercept 1 (kg/m^2^) | | | |
| --- | --- | --- | --- | --- | --- | --- | --- | --- | --- | --- | --- | --- | --- | --- | --- | --- | --- |
| Outcome | Metabolite  (SD) | value | 95%CI | | p value | value | 95%CI | | p value | value | 95%CI | | p value | value | 95%CI | | p value |
|  |  |  | LB | UB |  |  | LB | UB |  |  | LB | UB |  |  | LB | UB |  |
| **Men** |  |  |  |  |  |  |  |  |  |  |  |  |  |  |  |  |  |
| BMI at ~22  (kg/m2) | FBPALD.B | 13.82 | 11.17 | 16.47 | <2.2  e-16 | 0.20 | -1.44 | 1.84 | 0.83 | 0.03 | -2.24 | 2.31 | 0.93 | 0.93 | 0.81 | 1.05 | <2.2  e-16 |
| BMI at 37  (kg/m2) | FBPALD.B | 7.54 | 3.28 | 12.03 | 7.35  e-04 | 0.22 | -2.59 | 3.12 | 0.85 | 0.12 | -3.85 | 3.95 | 0.91 | 1.01 | 0.79 | 1.24 | 4.21  e-15 |
| BMI at 37  (kg/m2) | Glucose | 8.52 | 4.56 | 12.59 | 4.70  e-05 | 1.58 | -1.04 | 4.24 | 0.22 | -1.47 | -5.26 | 2.25 | 0.44 | 0.96 | 0.74 | 1.19 | 5.27  e-14 |
| **Women** |  |  |  |  |  |  |  |  |  |  |  |  |  |  |  |  |  |
| BMI at ~22  (kg/m2) | Glucose | 15.24 | 12.64 | 17.84 | <2.2  e-16 | -0.96 | -2.38 | 0.47 | 0.22 | 2.37 | -0.08 | 4.81 | 0.06 | 0.87 | 0.72 | 1.00 | <2.2  e-16 |
| BMI at ~22  (kg/m2) | Citrate | 15.30 | 12.75 | 17.86 | <2.2  e-16 | 0.21 | -1.25 | 1.66 | 0.71 | -1.26 | -3.67 | 1.15 | 0.3 | 0.87 | 0.72 | 1.01 | <2.2  e-16 |
| BMI at ~22  (kg/m2) | Pyruvate | 15.93 | 13.43 | 18.44 | <2.2  e-16 | 1.09 | -0.32 | 2.51 | 0.11 | -1.01 | -3.34 | 1.33 | 0.4 | 0.91 | 0.77 | 1.05 | <2.2  e-16 |
| BMI at 37  (kg/m2) | Citrate | 11.59 | 7.72 | 15.47 | 1.84  e-08 | -1.50 | -3.72 | 0.74 | 0.24 | 1.31 | -2.40 | 4.99 | 0.5 | 1.05 | 0.83 | 1.26 | <2.2  e-16 |
